# Effects of temperature settings on information quality of ChatGPT-3.5 responses: A prospective, single-blind, observational cohort study

**DOI:** 10.1101/2024.06.11.24308759

**Authors:** Akihiko Akamine, Daisuke Hayashi, Atsushi Tomizawa, Yuya Nagasaki, Chikae Akamine, Takahiro Fukawa, Iori Hirosawa, Orie Saigo, Misa Hayashi, Mitsuru Nanaoya, Yuka Odate

## Abstract

**Objective:** The effect of temperature settings on the quality of ChatGPT version 3.5 (OpenAI) responses related to drug information remains unclear. We investigated ChatGPT-3.5’s response quality on apixaban information with and without the temperature being set to 0.

**Methods:** On 6 September 2023, 37 questions regarding apixaban, derived from the frequently asked questions on the Bristol–Myers Squibb’s website, were entered into ChatGPT in Japanese. The primary endpoint was the effect of temperature settings on ChatGPT-3.5’s responses to apixaban-related questions. The response accuracy, clarity, detail, and adequacy were rated on a 5-point Likert scale by 10 pharmacists, with higher scores indicating higher response quality. Cumulative score means were analyzed using the Mann–Whitney U test. In the subgroup analysis, evaluators were limited to pharmacists at university hospitals. Welch’s t-test was employed in sensitivity analysis to validate primary endpoint findings.

**Results:** The mean scores for ChatGPT-3.5’s apixaban-related responses with (13.08) and without (14.40) the temperature being set to 0 were not significantly different (p = 0.064). Accuracy differed significantly (3.15 vs. 3.54, p = 0.045), whereas clarity, detail, and appropriateness were similar. Subgroup analysis (13.30 vs. 14.21, p = 0.394) and sensitivity analysis confirmed similar results (13.45 vs. 14.52, p = 0.105).

**Conclusions:** ChatGPT-3.5 temperature setting does not significantly affect overall responses to apixaban-related inquiries. However, the variance in accuracy suggests that ChatGPT-3.5 is unable to consistently provide precise responses. Hence, it is more suitable as a supplementary tool rather than a primary medical resource.

## Introduction

Recent advances in artificial intelligence (AI) have led to the development of sophisticated tools, such as ChatGPT [1], which are increasingly utilized in various fields, including pharmaceutical information services. ChatGPT, developed by OpenAI, has the potential to enhance patient care in the medical field by providing accurate information. Its efficacy in predicting drug–drug interactions highlights its important role in healthcare [2]. Furthermore, AI integration into medical safety education, including drug information services, is being actively explored. These investigations focus on addressing ethical and security issues associated with AI integration, ultimately aiming to provide comprehensive and personalized medical services [3]. In life sciences, AI has facilitated advances in research methods, protocols, and data analysis, enabling medical providers to make more effective decisions [4].

Despite these advances, the application of ChatGPT in drug information services remains challenging. In a previous real-world study, ChatGPT answered the majority of drug-related questions incorrectly or only partly correctly, highlighting the limitations of applying AI in the drug information field due to issues such as inaccurate content and a lack of references [5–9]. This performance inconsistency raises concerns regarding the reliability and robustness of AI-generated drug information. It is necessary to assess the accuracy and reliability of ChatGPT in providing pharmaceutical information, particularly under different operational settings, such as temperature, which can influence the model’s response style and content. A method for adjusting ChatGPT’s response quality by setting the temperature has been reported. ChatGPT temperature is a parameter that controls the diversity of the generated text and can be specified as a value between 0 and 1. At low temperatures, the generated text is more predictable and monotonous, whereas at high temperatures, the generated text may contain more diverse and random words and expressions [10].

The following limitations have been associated with previous studies: (i) a small number of ChatGPT response raters (<10) may have biased the ratings; (ii) drug information validation was lacking; (iii) responses with different temperature settings were not validated; and (iv) despite the evaluation of the response accuracy of ChatGPT, the clarity, detail, and appropriateness of the responses were not verified. Addressing these limitations would facilitate a more comprehensive evaluation of the quality of ChatGPT responses to pharmaceutical information.

This study compared and verified the quality of the answers provided by ChatGPT-3.5 regarding drug-related questions with and without a temperature setting of 0. The aim of this study was to evaluate the effect of temperature settings on the accuracy, clarity, detail, and appropriateness of ChatGPT-3.5 responses and to elucidate its reliability as a drug information tool. The drug of interest was oral apixaban (tablet; Eliquis®; Bristol–Myers Squibb, New York, NY, USA), the highest-selling oral drug in FY2022, excluding COVID-19 prophylaxis treatments and therapeutic modalities [11].

## Methods

### Ethics approval

This was an observational study. The Institutional Review Board for Observation and Epidemiological Study at the Doujin Hospital confirmed that no ethics approval was required (Date: 20 May 2023).

### Consent to participate

Informed consent was not required for this study because it did not involve human subjects.

### Study design

This prospective, single-blind, observational cohort study was conducted at eight hospitals, pharmacies, and pharmacy schools in Japan (Level of Evidence IV). Ten evaluators (D.H., A.T., Y.N., C.A., F.T., I.H., M.H., M.N., O.S., and Y.O.) participated in this study; among them, three pharmacists specialized in cancer, heart failure, and perioperative patient management. Data collection, creation of questions for ChatGPT, and analysis of ChatGPT responses were conducted between 1^st^ July and 30^th^ November 2023.

### Eligibility criteria of the pharmacists

We included pharmacists with at least 3 years of experience in hospital or pharmacy practice, who were employed in a facility utilizing apixaban. We excluded pharmacists who, according to the principal researcher, were deemed incapable of adequately evaluating drug information as well as pharmacists who used ChatGPT to obtain content relevant to the study during the evaluation period.

### Creating questions for ChatGPT

The principal researcher developed a total of 37 questions dissecting 35 frequently asked questions regarding apixaban posted in Japanese on the Bristol–Myers Squibb’s website, separating combined questions into individual queries, if necessary [12]. Three core researchers (D.H., A.T., and Y.N.) evaluated and approved the questions. In cases of disagreement, inputs were obtained from a fourth researcher (C.A.), and the principal investigator made the final decision.

### Evaluation of ChatGPT’s responses

On 6 September 2023, the primary researcher posted questions to ChatGPT-3.5 in Japanese. To maintain novelty, each question was asked from the same account using a “New Chat.” To maintain the temperature setting of ChatGPT at 0, we noted “Please use temperature of 0” at the end of each question. The reproducibility of responses was not considered in this study, and the first response obtained was evaluated. All questions required written responses, and no multiple-choice questions were included. All the textual prompts are provided in S1 File. The principal researcher used a Google form with one question that did not specify the temperature setting and two options (set and not set). These responses were randomized, blinded, and presented to the evaluators.

The evaluators were provided with answers with and without the temperature setting, in a random order. They were instructed to read the questions and their answers prior to the evaluation. Evaluators were asked to rate 296 responses using a 5-point Likert scale using four dimensions, (accuracy, clarity, detail, and appropriateness) [13]. Each question was answered only once, and the response options were rated on a scale of 1–5, with higher numbers indicating higher quality. The evaluation criteria are presented in S2 File. Evaluators consulted reliable sources of apixaban drug information (e.g., package inserts, interview forms, and Bristol–Myers Squibb’s website), as needed.

### Main analysis

The primary endpoint of the study was the quality of ChatGPT-3.5’s answers to apixaban-related questions with and without the temperature being set to 0. The accuracy, clarity, detail, and appropriateness of the responses were rated on a 5-point Likert scale (1–5), and the mean scores for all questions (4–20) were analyzed using Mann–Whitney U test. In addition, as secondary endpoints, the accuracy, clarity, detail, and appropriateness of ChatGPT-3.5’s answers were individually rated on a 5-point Likert scale (1–5) with and without a temperature setting of 0. The mean score for each category for all questions was analyzed using Mann–Whitney U test.

### Subgroup analysis

To confirm the robustness of the primary outcomes, we conducted a subgroup analysis that included only pharmacists affiliated with university hospitals, to evaluate the influence of affiliations of pharmacists on outcomes similar to the main analysis. University hospitals have reported higher patient satisfaction than general hospitals [14], but no difference in mortality or readmission rates by disease has been noted. However, since no studies have compared the quality of pharmacists at different institutions, only pharmacists from university hospitals were included in this analysis.

### Sensitivity analysis

To determine the effect of the statistical analysis method on the primary outcome, a sensitivity analysis was performed by changing the statistical analysis method to Welch’s t-test.

### Statistical analysis

The Shapiro–Wilk test was used to test the normality of the distribution of the age and career data of the participating pharmacists as continuous variables. Continuous variables were expressed as medians and means, whereas categorical data were expressed as absolute values and percentages. Welch’s t-test was used to analyze the means of continuous variables, and Mann–Whitney U test was used to analyze the medians [15,16]. Pharmacists with missing study data were excluded from the univariate analyses. However, when ≥20% of the data were missing, multiple imputations were planned using chained equations to create 100 sets of corresponding data. All statistical analyses were performed using the EZR version 1.36 software (Saitama Medical Center, Jichi Medical University, Saitama, Japan) [17]. All the tests were two-tailed. Statistical significance was set at p < 0.050. As this was an exploratory study, the sample size was not calculated. Nominal *p*-values were used to account for the multiplicity of analyses.

## Results

### Characteristics of pharmacists

Ten pharmacists evaluated all the responses, and none of the evaluators met the exclusion criteria. The age (p = 0.649) and career length (p = 0.551) of the pharmacists showed normal distributions. Pharmacist characteristics are listed in Table 1.

**Table 1.** Pharmacist characteristics evaluated at baseline (n = 10).

| Characteristic | Total<br>(n = 10) |
| --- | --- |
| Age |  |
| Median (IQR), years | 38.5 (32.8–42.8) |
| Mean (SD), years | 38.2 (6.3) |
| Sex, No. (%) |  |
| Male | 5 (50.0) |
| Female | 5 (50.0) |
| Career as a pharmacist |  |
| Median (IQR), years | 14.5 (9.0–20.3) |
| Mean (SD), years | 14.6 (6.7) |
| Academic history, No. (%) |  |
| Doctor | 1 (10.0) |
| Master | 4 (40.0) |
| Bachelor | 5 (50.0) |
| Affiliation |  |
| University hospital | 6 (60.0) |
| General hospital | 2 (20.0) |
| Pharmacy | 1 (10.0) |
| Faculty | 1 (10.0) |

Abbreviations: IQR, interquartile range; SD, standard deviation

### Primary outcome

All ChatGPT responses were in Japanese, eliminating the need to translate the responses. With the temperature set to 0, the median of the answers was 13.08 (interquartile range: 12.50–14.03), whereas it was 14.40 (interquartile range: 13.84–15.32) without a temperature setting, demonstrating no significant differences (p = 0.064). Answers with the temperature set at 0 had a lower rate of total scores of ≥16 (maximum: 20) than those without a temperature setting (7/37 [18.92%] vs. 17/37 [45.95%]; Fisher’s exact test, p = 0.024). The results of Mann–Whitney U test showed a significant difference between the mean scores for answers with and without the temperature being set to 0 (accuracy: 3.15 [interquartile range: 3.06–3.30] and 3.54 [interquartile range: 3.29–3.65]; p = 0.045). However, clarity, detail, and adequacy of answers were similar between groups (Table 2).

**Table 2.**
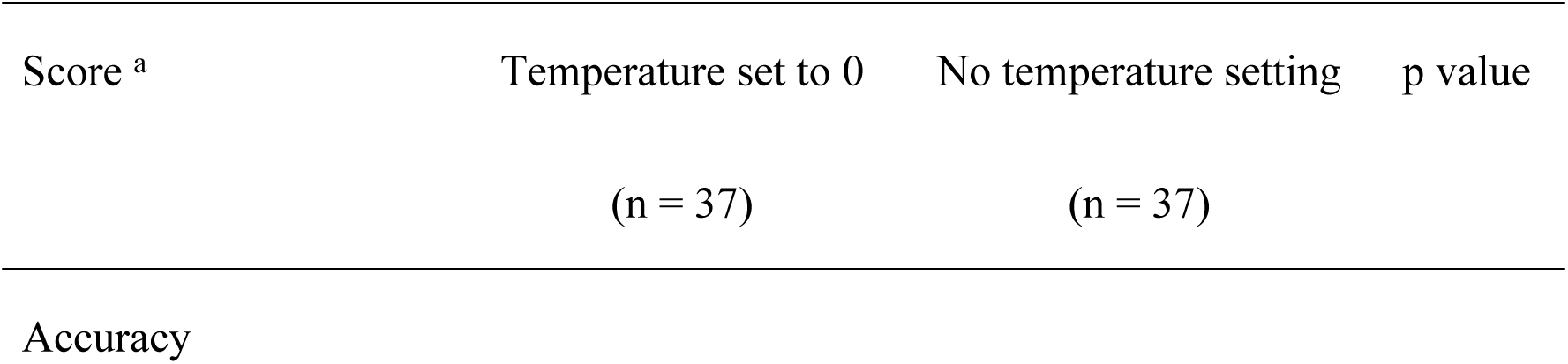
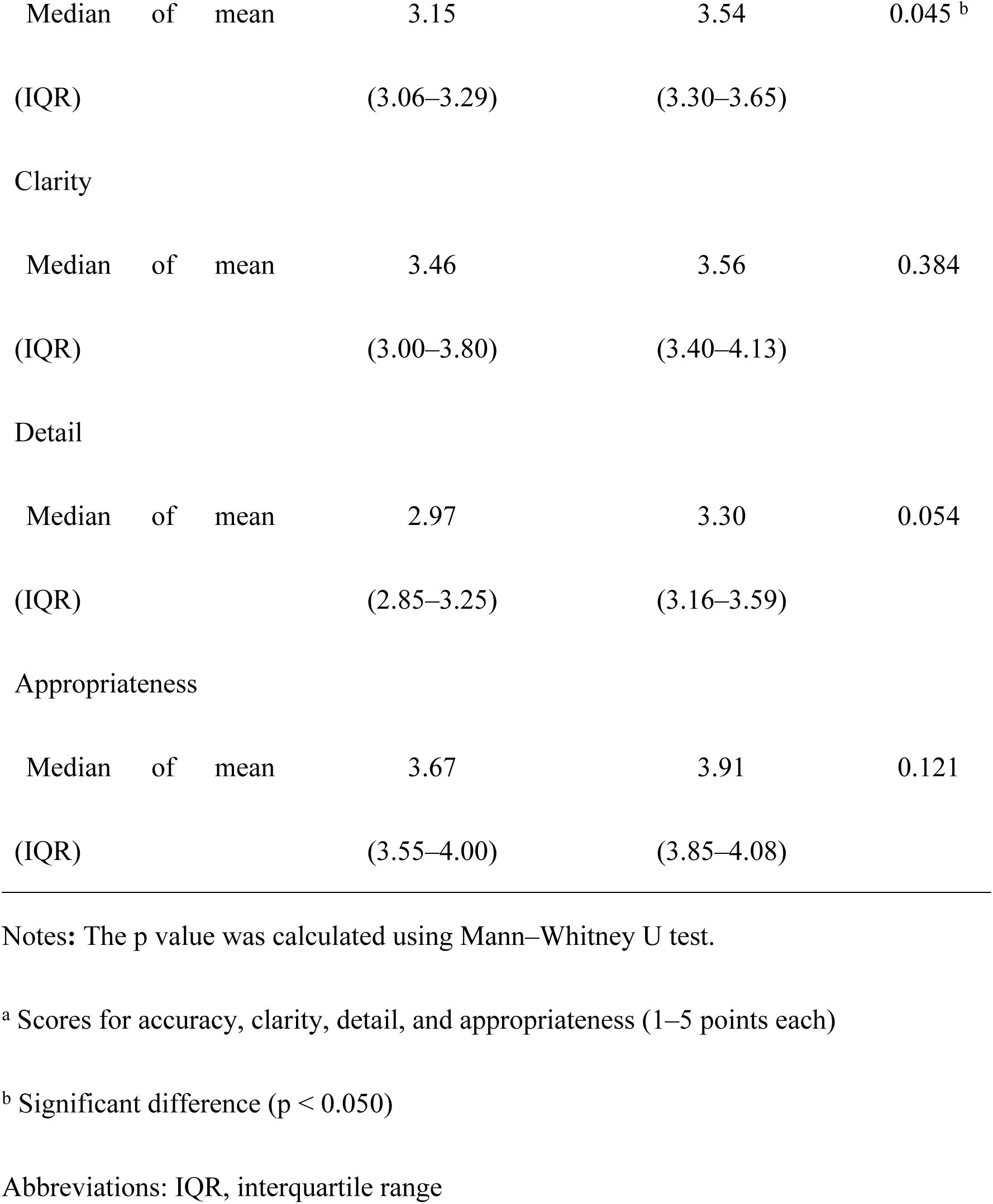
Comparisons of the scores of each endpoint for questions on apixaban drug information with and without the ChatGPT temperature setting (n = 74).

### Results of the subgroup analysis

The university hospital pharmacist subgroup analysis yielded median values of 13.30 (interquartile range; 12.00–15.02) and 14.21 (interquartile range: 13.45–15.32) for answers with and without the temperature being set at 0, respectively, demonstrating no significant differences (p = 0.394). Hence, the subgroup analysis yielded results similar to those of the primary analysis.

### Results of the sensitivity analysis

When the primary analysis method was revised to Welch’s t-test, the mean scores of the answers with and without the temperature being set to 0 were 13.45 (standard deviation; 1.51) and 14.52 (standard deviation; 1.28), respectively, which were not significantly different (p = 0.105). Thus, the sensitivity analysis yielded the same results as those of the primary analysis.

## Discussion

### Summary of key findings

This study yielded two important findings. First, the overall quality of ChatGPT-3.5’s responses in terms of accuracy, clarity, detail, and adequacy was consistent, regardless of the temperature setting, as evidenced by similar results across the primary endpoints, subgroup analyses, and sensitivity analyses. Second, responses with a temperature setting of 0 were less likely to have a total score ≥16 than those with no temperature setting (18.92% vs. 45.95%, Fisher’s exact test, p = 0.024). These findings provide a basis for further discussion on the implications of temperature settings on AI-generated drug information.

Although a previous study has shown that AI-based chatbots, including this version of ChatGPT, have robust search and information integration capabilities, particularly in clinical pharmacy [18], our study found a lower percentage of high-quality responses when the temperature was set to 0 than those without temperature settings. This finding is particularly interesting because it suggests a subtle effect of temperature setting on response quality, which has not been previously explored. Furthermore, it underscores the life-threatening consequences of using medication based on incorrect information. Thus, addressing and resolving this issue promptly is crucial. Additionally, users should ask detailed questions because the quality of ChatGPT answers depends on the phrasing of the questions.

### Strengths and weaknesses

This study has several strengths. It contributes markedly to the field of clinical pharmacy and AI-based tools as it provides unique insights into the impact of temperature setting on the quality of pharmaceutical information provided by ChatGPT-3.5. This specific focus on temperature settings and their influence on AI response quality has not been extensively explored in previous research. Second, the study employed a robust methodology, including a clear primary endpoint and comprehensive statistical analyses.

In addition, the study included 10 pharmacists in the evaluation, whose diversity provided a broader perspective, thereby reducing bias and enhancing the representativeness of the ratings. During the evaluation, the evaluators were not informed about whether the responses from ChatGPT-3.5 were temperature-adjusted, thus reducing potential order bias. Subgroup and sensitivity analyses were performed to ensure the consistency of results for the primary endpoints, thereby enhancing result reliability. This enabled us to generalize the results across different settings and rater profiles. Moreover, this study acknowledges ChatGPT-3.5’s limitations, particularly its lack of internet search capabilities and reliance on preexisting datasets. Our critical evaluation highlights the importance of continuous updates and improvements in AI tools to ensure their effective use in healthcare settings, particularly in domains where current and accurate information, such as drug data, is crucial. Finally, this study focused on a specific drug, apixaban, allowing for a detailed and focused analysis of AI performance in providing medication information. Although this approach limits the generalizability of the findings, it enables a more in-depth understanding of AI capabilities and limitations in the context of a single, widely used medication.

The study limitations must also be acknowledged. First, evaluators were limited to pharmacists, primarily those working at university hospitals. This specific professional background may have influenced the perception and evaluation of AI-generated responses. Second, we used the Japanese version of ChatGPT-3.5, and the results may vary for other languages due to differences in language processing and available datasets in the AI model.

### Interpretation

Consistent with our findings, the limitations of AI chatbots in effectively handling complex medical information have been highlighted in previous studies that have cited a lack of medicine-specific datasets and challenges in advanced reasoning [19]. Temperature settings designed to control response randomness may inadvertently affect the chatbots’ ability to access and integrate complex medical information effectively.

Although no significant difference was detected in the overall quality of responses from ChatGPT-3.5 across temperature settings, a lower percentage of high-quality responses was observed when the temperature was set to 0, thereby warranting further investigation. This emphasizes the importance of careful consideration of the AI chatbot settings in clinical applications and settings, ensuring they are optimized to provide accurate and relevant information.

The subgroup and sensitivity analyses conducted in our study provided additional insights into the robustness of ChatGPT-3.5’s responses to pharmaceutical inquiries. In the subgroup analysis limited to university hospital pharmacists, our findings remained consistent with the primary outcome. This consistency across different groups of raters reinforces the quality of ChatGPT-3.5 responses in a professional academic setting.

Furthermore, the sensitivity analysis performed using a different statistical test also supported the primary findings, showing non-significant differences in response quality. This methodological robustness enhances the credibility of our results, suggesting that the observed variance in accuracy is not a statistical anomaly but a characteristic of the AI model’s performance. However, the slight variance in accuracy observed in the primary analysis remains a matter of concern. Although this variance is not statistically significant, it could have implications in clinical settings where precise drug information is crucial. Previous studies have also indicated variability in AI responses in clinical scenarios, suggesting the need for the cautious application and continuous monitoring of AI tools in healthcare [2,20].

ChatGPT-3.5 does not have an internet search capability, constricting its ability to provide responses integrating the latest information, which is crucial in the drug information field. For example, although the package insert recommends the administration of Ondexxya in the event of life-threatening or difficult-to-staunch bleeding when consuming apixaban, no responses related to the administration of andexanet alfa (injection) (Ondexxya®; AstraZeneca, London, UK) were noted. This could be attributed to the fact that Ondexxya was not available in Japan until May 2022, and the ChatGPT data only extended until September 2021. This limitation is particularly relevant in clinical pharmacy practice where accurate and up-to-date information is paramount for patient safety. ChatGPT-3.5, however, contains limited learning data, which is a crucial factor to be considered in the field of drug information. Our findings underscore the importance of regularly updating and improving AI chatbots for their effective utilization in clinical pharmacies and healthcare settings.

### Future research

Although we found no significant difference in the overall quality of responses from ChatGPT-3.5 across temperature settings, a lower percentage of high-quality responses was observed when the temperature was set to 0, suggesting the need for further investigation and careful consideration of AI chatbot settings in clinical applications. Settings that optimize the information accuracy and relevance should be provided. Future studies should focus on the inclusion of various drugs to obtain a deeper understanding of the capabilities and limitations of ChatGPT-3.5 in relation to various drug classes and their respective complexities.

Additionally, the participation of a diverse group of healthcare professionals is essential for future evaluations to obtain a broader perspective on AI performance and its utility across the healthcare ecosystem. Given the global applicability of AI tools, it is critical to conduct similar studies in various linguistic and cultural contexts. This approach will aid in understanding the impact of language processing and cultural nuances on ChatGPT-3.5, aiming to evaluate the global validity and reliability of this tool. Further investigation is also required to determine the reasons for the variation in AI response quality, particularly under different temperature settings. Understanding the mechanisms that lead to this variability will guide the development of more consistent and reliable AI tools for clinical use.

Research focusing on the impact of AI tools on patient safety is critical. Finally, studies should examine the ethical and legal aspects of AI in healthcare, particularly regarding privacy, data security, and liabilities. Understanding these implications is essential for the responsible incorporation of AI tools into clinical practice.

## Conclusions

The use of temperature settings of ChatGPT-3.5 did not result in significant differences in the overall quality of responses to drug queries, specifically those related to apixaban. This suggests that ChatGPT-3.5 responses are not significantly affected by this setting. However, the variability in accuracy highlights the need for careful consideration when using this tool in clinical settings. Despite its potential as a supportive tool in pharmaceutical information retrieval, its limitations, including the lack of real-time internet access and the potential for the use of outdated information, must be acknowledged. Healthcare professionals should use ChatGPT-3.5 as a supplementary source, always verifying its output against current, evidence-based medical literature. Future research should aim to evaluate chatbot performance across a broader range of medications with larger and more diverse groups of healthcare professionals for a comprehensive understanding of the capabilities and limitations of chatbots in the context of clinical pharmacy.

## Data Availability

datasets generated and/or analyzed during the current study are available in the Mendeley repository, https://data.mendeley.com/datasets/dw2993xm2n/1

## Acknowledgments

We would like to thank Hiroki Yoshida for advice on statistical analysis and Editage (www.editage.com) for English language editing and journal submission support. The authors have authorized the submission of this manuscript through Editage.

## Supporting information

**S1 File. Questions entered in Japanese were translated into English for this study. S2 File. Criteria for evaluating the quality of responses used in this study.**

